# Trends in demographic and clinical characteristics of patients enrolled in HIV care and subsequent Antiretroviral Therapy initiation in the Central Africa International epidemiology Database to Evaluate AIDS (CA-IeDEA) Adult Cohort 2004-2018

**DOI:** 10.1101/2020.07.17.20156141

**Authors:** Adebola A. Adedimeji, Donald R. Hoover, Qiuhu Shi, Hae-Young Kim, Ellen Brazier, Jonathan Ross, Gad Murenzi, Christella Twizere, Patricia Lelo, Dominique Nsonde, Rogers Ajeh, Anastase Dzudie, Denis Nash, Marcel Yotebieng, Kathryn M. Anastos, for the Central Africa IeDEA Consortium

**Affiliations:** Department of Epidemiology and Population Health, Albert Einstein College of Medicine/Montefiore Medical Center, Bronx, NY; Department of Statistics, Rutgers University, Piscataway, NJ; Department of Public Health, School of Health Sciences and Practice, New York Medical College, Valhalla, NY USA; School of Public Health, City University of New York, New York, NY; Division of General Internal Medicine, Montefiore Medical Center, Bronx, NY; Division of Clinical Education, Rwanda Military Hospital, Kanombe, Kigali, Rwanda; Centre National de Reference en Matière de VIH/SIDA, Burundi; Pediatric Hospital Kalembe Lembe, Lingwala, Kinshasa, Democratic Republic of Congo; CTA Brazzaville, Republic of Congo; Clinical Research Education, Networking and Consultancy, Yaounde, Cameroon

## Abstract

**Background:** The Central Africa International epidemiology Database to Evaluate AIDS (CA-IeDEA) is a prospective study investigating impact, progression and long-term outcomes of HIV/AIDS among people living with HIV (PLWH) in Burundi, Cameroon, Democratic Republic of Congo (DRC), Republic of Congo (ROC) and Rwanda. We described trends in baseline demographic, clinical and immunological characteristics of patients aged >15 years entering into HIV care in the participating CA-IeDEA site and subsequent ART initiation.

**Materials and Methods:** Information on socio-demographic characteristics, height, weight, body mass index (BMI), CD4 cell count, WHO staging and ART status at entry into care from 2004 to 2018 were extracted from clinic records of patients aged >15 years enrolling in HIV care at participating clinics in Burundi, Cameroon, DRC, ROC and Rwanda. We assessed trends in patient characteristics at enrollment in HIV care and ART initiation at the participating site and calculated proportions, means and medians (interquartile ranges) for the main variables of interest.

**Results:** Among 69,176 participants in the CA-IeDEA cohort, 39% % were from Rwanda,, 24% from ROC, 18% from Cameroon, 14% from Burundi and 5% from DRC. More women (66%) than men were enrolled in care and subsequently initiated ART. Women were also younger (32 years) than men (38 years) (p= <0.001) when they enrolled in care or subsequently initiated ART at the participating site. Trends over time show increases in median CD4 cell count of 190 cells/uL in 2004 to 334 cells/uL in 2018 at enrollment. Among those with complete data on CD4 counts (60%), women had higher median CD4 cell count at care entry in the CA-IeDEA site (299 cells/uL) versus men (249 cells/uL; p= < 0.001).Trends in proportion of patients using ART show an increase from 16% in 2004 to 75% in 2018 among those initiating ART for the first time after entry into care in the participating site. As expected, median CD4 generally increased after ART initiation (p= <0.001).

**Conclusion:** Trends from 2004-2018 in the characteristics of patients participating in the CA-IeDEA cohort highlight improvements over time at entry into care and subsequent ART in all participating sites.

## Introduction

Globally, over 37.9 million people aged >15 years are living with HIV of whom 24.5 million were estimated to be accessing antiretroviral therapy (ART).^**1**^ Expanded access to ART contributed to improved quality of life and declines in AIDS-related mortality in people living with HIV (PLWH). To consolidate these gains and end the threat of HIV to global public health, the Joint United Nations Program on AIDS (UNAIDS) recommended the “90-90-90” strategy to ensure 90% of PLWH are diagnosed and know their status, 90% of those diagnosed are initiated on ART and 90% of those initiating ART attain viral suppression. To facilitate the attainment of these global targets, the World Health Organization (WHO) put out the “Treat All” guidelines, which recommended immediate ART initiation for all persons diagnosed with HIV regardless of CD4 or viral load.^**2**^

Despite expanded ART access, sub-Saharan Africa (SSA) is still disproportionately burdened by HIV. For instance in 2018, 73% of all new HIV infections diagnosed globally were in the region.^**3**^ In most of SSA, progress towards the 90-90-90 targets is uneven with many countries in East and Southern Africa making remarkable progress compared with those in West and Central Africa.^**4, 5**^ Whereas, the 25 countries in West and Central Africa comprised only 6% of the world’s population, they accounted for 18% of global HIV burden.^**6**^ The UNAIDS estimated that 4.7 million people living with HIV in West and Central Africa lack access to treatment, a sharp contrast with treatment coverage in East and Southern Africa regions.^**6**^ These discrepancies led to a regional catch up plan to accelerate progress in West and Central Africa, but this has not translated into meaningful gains in scaling up access for millions of PLWH in the region. ^**6**^

The lack of progress reflects disparities in access to care for many PLWH in Central Africa who continue to experience late diagnosis, non-linkage to care and poor HIV outcomes.^**1,2,3,4,5,6,7**^ Social and structural instability, especially stigma, weak health systems, civil unrest and other contextual factors impeded access to HIV care and progress towards universal targets.^**5-8**^ To address gaps in access to care for PLWH in Central Africa, it is critical to understand the characteristics of PLWH enrolling in care at given sites and subsequently initiating ART. Additionally, knowledge of trends in demographic, immunological and clinical characteristics among PLWH enrolled in care can highlight challenges in implementing effective strategies to facilitate early diagnosis and linkage to care for PLWH in these countries.

The Central Africa International epidemiology Database to Evaluate AIDS (CA-IeDEA) research consortium was established with funding from the United States National Institutes of Health to be a resource to investigate the impact, progression and long-term outcomes of the HIV/AIDS epidemic in Central Africa and answer epidemiologic research questions that cannot be answered by separate individual cohorts. The CA-IeDEA routinely collects socio-demographic, clinical and immunologic data from patients enrolled in HIV care at participating treatment sites in Burundi, Cameroon, Democratic Republic of Congo, Republic of Congo and Rwanda. These data provide insights on trends in sociodemographic, clinical and immunological characteristics of adult patients entering HIV care in the given site and subsequently initiating ART in the participating countries. The objective of this study was to examine these trends in sociodemographic, clinical and immunological characteristics of adult patients aged >15 years entering care in the current site and subsequently initiating ART in CA-IeDEA regional cohort from 2004-2018 with the goal of describing trends in these characteristics over time.

## Methods

### Study Settings

The Central Africa IeDEA (CA-IeDEA) is one of the 7 regional cohorts within the global IeDEA consortium [https://www.iedea.org/], with other cohorts in East Africa, Southern Africa, West Africa, North America, the Caribbean and Central/South America and Asia-Pacific regions. The overall IeDEA cohort, which has been previously described^**9**^ consists of 1.7 million PLWH of which 1.4 million in SSA are on ART.

The CA-IeDEA cohort currently comprises 19 active sites in Burundi, Cameroon, Democratic Republic of Congo (DRC), Republic of Congo (ROC), and Rwanda, and one historical site in DRC that contributed patient data to the cohort until mid 2013. The 19 active sites are predominantly public sector health facilities, including tertiary-level university teaching hospitals and primary level health centers. All the sites are located in urban or peri-urban settings. Details of CA-IeDEA sites have been previously described.^**9**^ Patients ever enrolling in HIV care at these participating sites are prospectively included in the CA-IeDEA cohort. Data from routine patient care are regularly extracted from patient records and electronic medical records and harmonized into a regional dataset for use in country-level and regional analyses. As of July 2018, there were approximately 70,000 patients in the CA-IeDEA database.

### Study Design and population

This analysis included all adult patients aged >15 years enrolled for HIV care in the CA-IeDEA cohort at participating sites in Burundi, Cameroon, Democratic Republic of Congo, Republic of Congo and Rwanda. Data obtained during routine patient care in each country are extracted from an electronic data collection and storage system in each country, for example, the Open Medical Records System (in Rwanda), SANTIA (in ROC), SIDA (in Burundi) and RedCap (in Cameroon and DRC) ^**10**^ as mandated by the local health authority in each country. These data are then uploaded to a central data server at the CA-IeDEA data center in New York where they undergo cleaning and harmonization prior to analysis.

### Ethical Approval

Ethical approval for the study was granted by the Albert Einstein College of Medicine Institutional Review Board in New York, and the relevant ethics review boards in Rwanda (Rwanda National Health Research Committee and the National Ethics Committee), Burundi (Comite National d’Ethique), Cameroon (Comite National D’Ethique la Recherche en la Sante Humaine-CNERSH) DRC (Ministere de l’Enseignement Superieur et Universitaire, University de Kinshasa Ecole de Sante Publique) and in the Republic of Congo (Comite d’Ethique De La Recherche En Sciences De La Sante (CERRSSA). Data for the Central Africa International Epidemiology to Evaluate AIDS (CA-IeDEA) is available upon request.

### Measures

The following socio-demographic characteristics were assessed at entry into care at the participating CA-IeDEA site: age, gender, and marital status, reported pregnancy among women and route of entry into HIV care in the participating site such as voluntary counseling and testing (VCT), prevention of mother-to-child transmission (PMTCT), Tuberculosis (TB) clinic or In-patient care. Clinical and immunological characteristics assessed at entry into HIV care at the participating CA-IeDEA site included weight, height, BMI, CD4 cell count, WHO stage at enrollment and ART usage. ART usage prior to enrollment was defined as ART use >30 days before entering care in the participating site. ART use immediately after entering HIV care at the participating site was defined as ART initiation starting <=30 days before enrollment to 30 days after enrollment. We considered CD4 cell count measures at enrollment defined as the closest CD4 measure available at enrollment date up to 3 months and 30 days prior to enrollment date. CD4 cell count measure at ART initiation was defined as first CD4 measure available 30 days and up to 3 months after ART initiation date among patients who initiated ART use after enrollment.

### Statistical Analysis

For this analysis, we calculated proportions, means (standard deviations) and medians (interquartile ranges) for the main variables of interest. We ultimately categorized all continuous variables as shown in the Tables and used Chi-Square tests to make comparison of all variables between groups. We also calculated the same data summaries by calendar year of enrollment into HIV care at the CA-IeDEA participating site and by country.

## Results

### Demographic and clinical profile of all adults at enrollment into care

Table 1 presents the socio-demographic and clinical profile of 69,176 patients entering HIV care in the CA-IeDEA cohort in all the five participating countries. The largest proportion of patients (39%) was in Rwanda and the smallest proportion (5%) was in the DRC. There were almost twice as many women (66%) as men enrolled in the participating sites. Most patients reported being married (28%) or single (23%). The median age at enrollment in the participating CA-IeDEA sites was 35 years (IQR: 28,42). Women were generally younger (33 years; IQR: 27, 40) than men (38 years; IQR: 32, 45) at enrollment in the participating clinics. The largest proportion of patients enrolled in HIV care at the participating site through voluntary counseling and testing (VCT) clinics (31%) and prevention of mother-to-child (PMTCT) clinics (9%).

**Table 1:** Demographic and clinical characteristics of all adult patients at enrollment into care by sex.

| Variable | Code | Total | Women | Men | P-value |
| --- | --- | --- | --- | --- | --- |
| Adult |  | 69176 | 45933 (66.4) | 23243 (33.6) |  |
| Pregnant |  |  | 2869 (6.2) |  |  |
| Age (yrs) median (IRQ) | Median (IQR) | 35(28,42) | 33(27,40) | 38(32,45) | <0.001 |
|  | 15-24 | 8894 (12.9) | 7291 (15.9) | 1603 (6.9) | <0.001 |
|  | 25-34 | 25623 (37.0) | 19088 (41.6) | 6535 (28.1) |  |
|  | 35-44 | 22031 (31.8) | 13113 (28.5) | 8918 (38.4) |  |
|  | 45+ | 12628 (18.3) | 6441 (14.0) | 6187 (26.6) |  |
| Point of entry into care | PMTCT | 6530 (9.4) | 5623 (12.2) | 907 (3.9) | <0.001 |
|  | TB | 38 (0.1) | 27 (0.1) | 11 (0.0) |  |
|  | VCT | 21285 (30.8) | 12372 (26.9) | 8913 (38.3) |  |
|  | Inpatient | 1009 (1.5) | 524 (1.1) | 485 (2.1) |  |
|  | Other | 6177 (8.9) | 3904 (8.5) | 2273 (9.8) |  |
|  | Unknown | 34137 (49.3) | 23483 (51.1) | 10654 (45.8) |  |
| Marital status | Single | 15894 (23.0) | 10911 (23.8) | 4983 (21.4) | <0.001 |
|  | Married/in union | 19345 (28.0) | 10735 (23.4) | 8610 (37.0) |  |
|  | Living with partner | 3413 (4.9) | 2372 (5.2) | 1041 (4.5) |  |
|  | Divorced | 3153 (4.6) | 2326 (5.1) | 827 (3.6) |  |
|  | Widowed | 5073 (7.3) | 4299 (9.4) | 774 (3.3) |  |
|  | Unknow/Missing | 22298 (32.2) | 15290 (33.3) | 7008 (30.2) |  |
| Weight for adults, Kg | Median (IQR) | 57(50,65) | 56(49,64) | 59(53,67) | <0.001 |
|  | Missing n (%) | 20702(29.9) | 13901(30.3) | 6801(29.3) |  |
| Height for adults, Cm | Median (IQR) | 163(158,169) | 160(155,165) | 170(165,175) | <0.001 |
|  | Missing n (%) | 20555(29.7) | 13551(29.5) | 7004(30.1) |  |
| BMI | Median (IQR) | 21.63(19.27,24.38) | 22.07(19.53,24.98) | 20.94(18.93,23.31) | <0.001 |
|  | Missing n (%) | 31156(45.0) | 20702(45.1) | 10454(45.0) |  |
| CD4 count (cells/uL) | Median (IQR) | 282(139,476) | 299(151,501) | 249(119,427) | <0.001 |
|  | <100 | 7500 (10.8) | 4610 (10.0) | 2890 (12.4) | <0.001 |
|  | 100-199 | 7513 (10.9) | 4846 (10.6) | 2667 (11.5) |  |
|  | 200-349 | 10093 (14.6) | 6739 (14.7) | 3354 (14.4) |  |
|  | >350 | 16456 (23.8) | 11811 (25.7) | 4645 (20.0) |  |
|  | Missing n (%) | 27614 (39.9) | 17927 (39.0) | 9687 (41.7) |  |
| WHO Stage at enrollment | Stage I | 21004 (30.4) | 14247 (31.0) | 6757 (29.1) | <0.001 |
|  | Stage II | 12961 (18.7) | 8659 (18.9) | 4302 (18.5) |  |
|  | Stage III | 11167 (16.1) | 6967 (15.2) | 4200 (18.1) |  |
|  | Stage IV | 3698 (5.3) | 2257 (4.9) | 1441 (6.2) |  |
|  | Missing n (%) | 20346 (29.4) | 13803 (30.1) | 6543 (28.2) |  |
| Country | Rwanda | 26930 (38.9) | 16651 (36.3) | 10279 (44.2) | <0.001 |
|  | Burundi | 9720 (14.1) | 6409 (14.0) | 3311 (14.2) |  |
|  | Cameroon | 12482 (18.0) | 8323 (18.1) | 4159 (17.9) |  |
|  | DRC | 3292 (4.8) | 2718 (5.9) | 574 (2.5) |  |
|  | ROC | 16752 (24.2) | 11832 (25.8) | 4920 (21.2) |  |
| Time Period | Year <= 2006 | 15054 (21.8) | 10246 (22.3) | 4808 (20.7) | <0.001 |
|  | Year 2007 - 2010 | 17488 (25.3) | 11717 (25.5) | 5771 (24.8) |  |
|  | Year 2011 - 2014 | 17761 (25.7) | 12125 (26.4) | 5636 (24.2) |  |
|  | Year >= 2015 | 18873 (27.3) | 11845 (25.8) | 7028 (30.2) |  |
| ART Status at enrollment | Not on ART | 32233 (46.6) | 21357 (46.5) | 10876 (46.8) | <0.001 |
|  | Before enrollment* | 7601 (11.0) | 5268 (11.5) | 2333 (10.0) |  |
|  | Immediately after enrollment** | 29342 (42.4) | 19308 (42.0) | 10034 (43.2) |  |

\*: >30 days prior to enrollment day, \*\* within 30 days after enrollment day <sup>1</sup> By Chi-Square Tests

The median weight, height and BMI for patients at entry into HIV care at participating clinics was 57 kg (IQR: 50,65); 163cm (IQR: 158,169) and 21.63 kg/m^2^ (IQR:19.27,24.38) respectively. Median CD4 at entry into care at the participating site was 282 cells/uL (IQR:139, 476) among those for whom the CD4 cell count measure was available. Women enrolled in care at the participating site with higher CD4 count (299 cells/uL, IQR: 151, 501) than men (249 cells/uL; IQR: 119, 427). About half of patients (49.9%) enrolled in care with WHO Stage I or Stage II disease. Slightly higher proportion of men (24%) enrolled in care with advanced HIV disease (WHO Stage III and Stage IV) than women (20%).

About 46% of patients who enrolled in care at the participating sites did not use any antiretroviral therapy (ART) up to 3 months post enrollment; 11% initiated ART prior to (> 30 days) enrollment in HIV care at the participating site, and 42% initiated ART immediately after enrollment in HIV care at the participating site. The proportions of those who initiated ART before enrollment in HIV care and after enrollment at the participating sites were similar among men and women.

### Trends in measures of body mass index, CD4 cell count, and WHO stage at enrollment into care in all participating clinics

Figure 1 show calendar year trends in having a recent measure of body mass index (BMI), CD4 cell count and WHO staging at enrollment in the participating CA-IeDEA site for patients in all participating countries and separately for each country. Overall, there was a steady increase in the proportion of patients with measures of BMI from 2004 to up to the period after 2017. There was a similar gradual increase in the proportion of patients with WHO staging from 2004-2007 before a gradual decrease from 2008 and a leveling off at 78% after 2017. A similar pattern emerged regarding availability of measures of CD4 cell count, which increased in all years, except in 2010 and 2014 when there were slight decreases and then a sharp decline in the period after 2017.

**Fig. 1:**
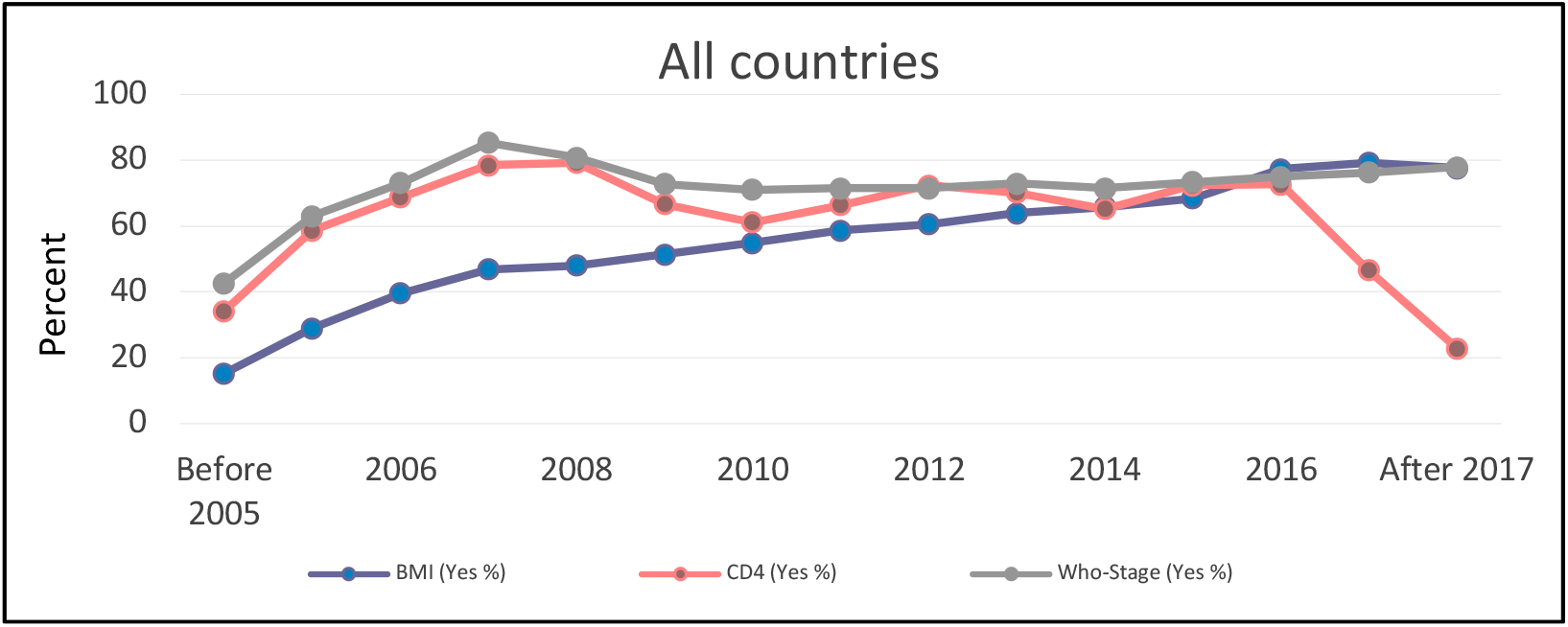
Trends measures of BMI, CD4 cell count and WHO stage at enrollment in all countries participating in the CA-IeDEA cohort.

Disaggregated data of trends in measures of BMI, CD4 cell count and WHO staging by calendar years in each participating country are shown in supplementary figures S1-S5.

Starting with Burundi (Figure S1) where measures were only available to this study from 2009 on the proportion of patients with BMI and WHO Stage measures increased between 2009 and 2014 before a sharp decline for BMI and leveling off for WHO stage between 2015 and 2017, and slight decline in the period after 2017. Trends in available CD4 cell count measures were declined from 2009 and 2015, rose slightly in 2016 when the Treat All guidelines ended the CD4 eligibility criterion for ART and thus declined again from 2017 on. In Cameroon (Figure S2) trends in measures of BMI show a gradual increase in the proportion of patients with this measure from 2004-2008, a gradual decrease until 2015 before a sharp increase since 2016. Trends in proportion of patients with measures of CD4 fluctuated between 2004 and 2016 before a sharp decline since 2017. The proportion of patients with measure of BMI was stable over time.

In the DRC, (Figure S3), trends in reported measures for BMI and WHO staging at enrollment in HIV care was available for nearly all patients except in 2014 when measures were available for about 84% of patients. Measures of CD4 cell count was available for at least 80% of patients until the year 2013 when there was a sharp decline that culminated in less than 1% of patients with measures of CD4 reported since 2017 when CD4 measures were no longer needed to determine eligibility for ART after 2016. In ROC (Figure S4) availability of measures of CD4, WHO stage and BMI rose between 2004 and 2007. From 2008 the proportion of patients with these measures continued to fluctuate. Trends in the availability of data for these measures tended upward since 2014 peaking at 83% for measures of BMI and CD4 and 45% for WHO stage.

Similar trends were observed in Rwanda (Figure S5) where the proportion of patients with measures of CD4 cell count and WHO stage remained high (i.e. >70%) and stable over time before a downward slope since 2016. The most significant decrease relates to the measure of CD4, which decreased to about one third of the patients in the year 2017 and beyond, which coincided with the implementation of the Treat All guidelines. Unlike measures of CD4 and WHO stage, measures of BMI, overall was available for less than 50% of patients, but rose steadily through the years, peaking at 80% in 2017.

### Characteristics of patients initiating ART within 30 days of enrolling in HIV care in the current in CA-IeDEA clinic

The characteristics of patients who first initiated ART within 30 days of enrollment in care at the participating clinic are shown in table 2. Overall, 29,342 participants were initiated on ART within 30 days of their enrollment of whom 66% were women. Median age among this first time ART initiators when they enrolled in care at the participating site was 35 years (IQR:29,43). Women were slightly younger than men (34 years vs 39 years) (p= <.0.001). Median CD4 among this first time ART initiators within 30 days of enrollment was was 203 cell/uL (IQR: 100,340). Median CD4 cell count at ART initiation was higher among women (216 cell/uL; IQR: 109, 356) than men (182 cell/uL; IQR: 82,309), p= < 0.001. About 44% of first time ART initiators did so in the year 2015 and beyond. Overall, 36% and 20% of first time ART initiators were missing CD4 cell count and WHO staging measures.

**Table 2:** Characteristics of patients initiating ART use within 30 days of enrollment into HIV care at participating CA-IeDEA clinic.

| Variable | Code | Total | Women | Men | P-value |
| --- | --- | --- | --- | --- | --- |
| Adult |  | 29342 | 19308 (65.8) | 10034 (34.2) |  |
| Pregnant |  |  | 1700 (8.8) |  |  |
| Age (yrs) median (IRQ) | Median (IQR) | 35(29,43) | 34(28,41) | 39(32,45) | <0.001 |
|  | 15-24 | 3402 (11.6) | 2747 (14.2) | 655 (6.5) | <0.001 |
|  | 25-34 | 10324 (35.2) | 7668 (39.7) | 2656 (26.5) |  |
|  | 35-44 | 9654 (32.9) | 5700 (29.5) | 3954 (39.4) |  |
|  | 45+ | 5962 (20.3) | 3193 (16.5) | 2769 (27.6) |  |
| Point of entry into care | PMTCT | 2748 (9.4) | 2479 (12.8) | 269 (2.7) | <0.001 |
|  | TB | 7 (0.0) | 4 (0.0) | 3 (0.0) |  |
|  | VCT | 7219 (24.6) | 3908 (20.2) | 3311 (33.0) |  |
|  | Inpatient | 434 (1.5) | 209 (1.1) | 225 (2.2) |  |
|  | Other | 2342 (8.0) | 1430 (7.4) | 912 (9.1) |  |
|  | Unknown | 16592 (56.5) | 11278 (58.4) | 5314 (53.0) |  |
| Marital status | Single | 8427 (28.7) | 5710 (29.6) | 2717 (27.1) | <0.001 |
|  | Married/in union | 9388 (32.0) | 5251 (27.2) | 4137 (41.2) |  |
|  | Living with partner | 2145 (7.3) | 1535 (8.0) | 610 (6.1) |  |
|  | Divorced | 1538 (5.2) | 1106 (5.7) | 432 (4.3) |  |
|  | Widowed | 2669 (9.1) | 2273 (11.8) | 396 (3.9) |  |
|  | Unknow/Missing | 5175 (17.6) | 3433 (17.8) | 1742 (17.4) |  |
| Weight for adults, Kg | Median (IQR) | 58(50,65) | 56(49,65) | 60(53,68) | <0.001 |
|  | Missing n (%) | 6254(21.3) | 4224(21.9) | 2030(20.2) |  |
| Height for adults, CM | Median (IQR) | 164(158,170) | 160(156,165) | 170(165,175) | <0.001 |
|  | Missing n (%) | 4943(16.8) | 3122(16.2) | 1821(18.1) |  |
| BMI | Median (IQR) | 21.55(19.10,24.34) | 21.91(19.27,24.89) | 20.96(18.82,23.42) | <0.001 |
|  | Missing n (%) | 9905(33.8) | 6500(33.7) | 3405(33.9) |  |
| CD4 count (cells/uL) | Median (IQR) | 203(100,340) | 216(109,356) | 182(82,309) | <0.001 |
|  | <100 | 4686 (16.0) | 2863 (14.8) | 1823 (18.2) | <0.001 |
|  | 100-199 | 4575 (15.6) | 2983 (15.4) | 1592 (15.9) |  |
|  | 200-349 | 5160 (17.6) | 3457 (17.9) | 1703 (17.0) |  |
|  | >350 | 4418 (15.1) | 3259 (16.9) | 1159 (11.6) |  |
|  | Missing | 10503 (35.8) | 6746 (34.9) | 3757 (37.4) |  |
| WHO Stage at enrollment | Stage I | 9171 (31.3) | 6160 (31.9) | 3011 (30.0) | <0.001 |
|  | Stage II | 6348 (21.6) | 4184 (21.7) | 2164 (21.6) |  |
|  | Stage III | 6021 (20.5) | 3721 (19.3) | 2300 (22.9) |  |
|  | Stage IV | 1935 (6.6) | 1180 (6.1) | 755 (7.5) |  |
|  | Missing | 5867 (20.0) | 4063 (21.0) | 1804 (18.0) |  |
| Country | Rwanda | 9091 (31.0) | 5289 (27.4) | 3802 (37.9) | <0.001 |
|  | Burundi | 3900 (13.3) | 2671 (13.8) | 1229 (12.2) |  |
|  | Cameroon | 9371 (31.9) | 6148 (31.8) | 3223 (32.1) |  |
|  | DRC | 1046 (3.6) | 847 (4.4) | 199 (2.0) |  |
|  | ROC | 5934 (20.2) | 4353 (22.5) | 1581 (15.8) |  |
| Time Period | Year <= 2006 | 3540 (12.1) | 2343 (12.1) | 1197 (11.9) | <0.001 |
|  | Year 2007 - 2010 | 5395 (18.4) | 3643 (18.9) | 1752 (17.5) |  |
|  | Year 2011 - 2014 | 7618 (26.0) | 5432 (28.1) | 2186 (21.8) |  |
|  | Year >= 2015 | 12789 (43.6) | 7890 (40.9) | 4899 (48.8) |  |

\* from 30 days prior to enrollment to 30 days after enrollment. <sup>1</sup> By Chi-Square Test

### Characteristics of patients already using ART prior to enrollment in HIV care at participating CA-IeDEA clinic

Table 3 shows the characteristics of 7, 601 patients who were already using ART prior to enrolling in HIV care in the current CA-IeDEA clinic. Presumably, these were patients transferring from a previous site where they were already initiated on ART after HIV diagnosis. Women made up about two-thirds (69%) of patients. Median age among ART experienced users was 36 years (IQR:30,43) and women were younger (35 years, IQR: 29, 41) than men (40 years IQR: 33, 47) (p= <0.001). Voluntary Counseling and Testing (38%) and PMTCT (11%) were the main sources of entry into care for ART experienced patients. ART experienced women had higher median CD4 cell/uL than men 392 (IQR 232,584) than ART experienced men 293 (IQR 160, 454). About 61% of experienced ART users were initiated on ART prior to 2015.

**Table 3:**
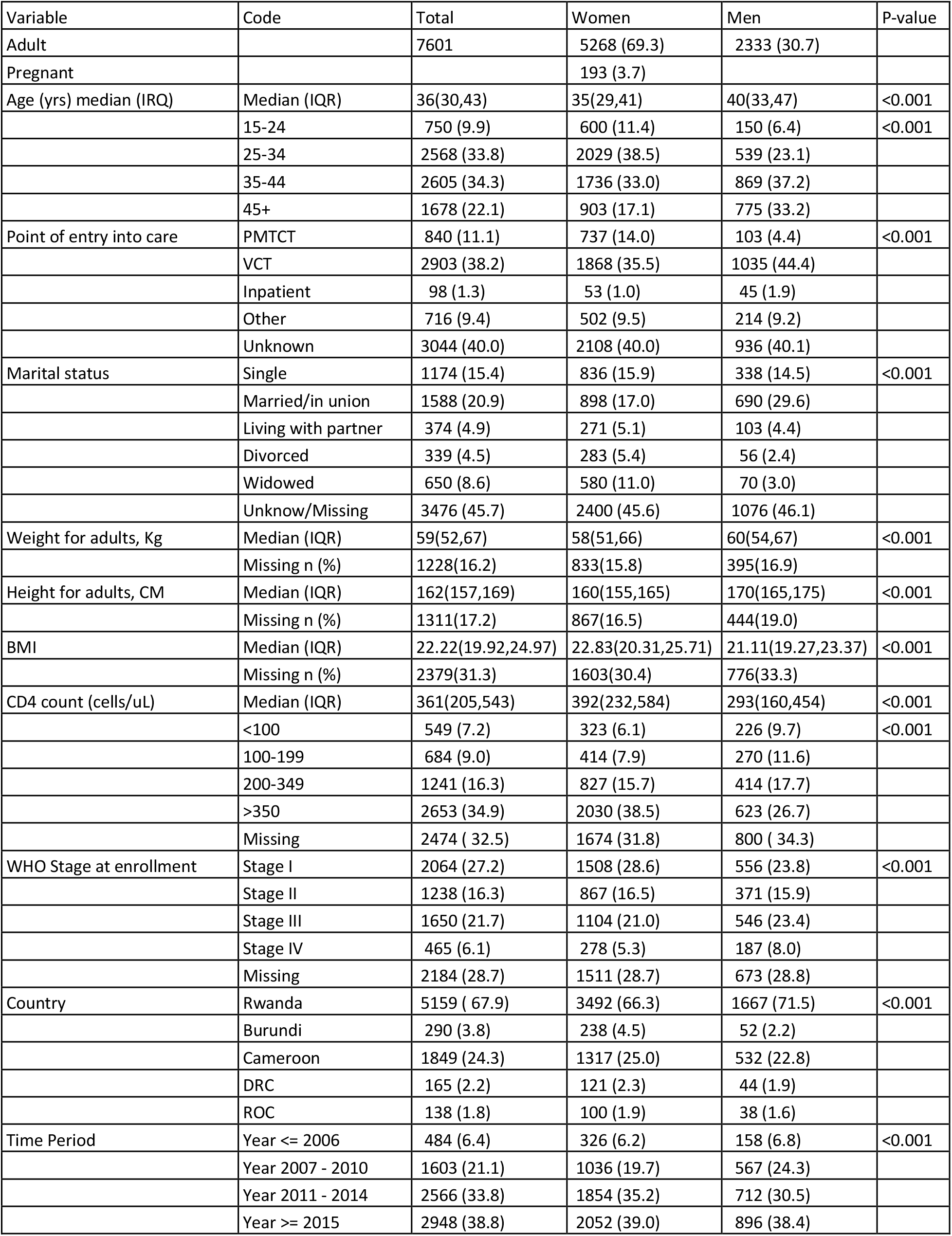
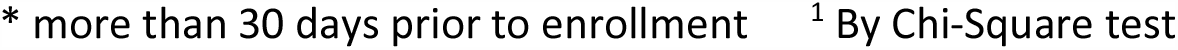
Characteristics of patients using ART prior to enrollment in HIV care at participating CA-IeDEA clinic.

| Variable | Code | Total | Women | Men | P-value |
| --- | --- | --- | --- | --- | --- |
| Adult |  | 7601 | 5268 (69.3) | 2333 (30.7) |  |
| Pregnant |  |  | 193 (3.7) |  |  |
| Age (yrs) median (IRQ) | Median (IQR) | 36(30,43) | 35(29,41) | 40(33,47) | <0.001 |
|  | 15-24 | 750 (9.9) | 600 (11.4) | 150 (6.4) | <0.001 |
|  | 25-34 | 2568 (33.8) | 2029 (38.5) | 539 (23.1) |  |
|  | 35-44 | 2605 (34.3) | 1736 (33.0) | 869 (37.2) |  |
|  | 45+ | 1678 (22.1) | 903 (17.1) | 775 (33.2) |  |
| Point of entry into care | PMTCT | 840 (11.1) | 737 (14.0) | 103 (4.4) | <0.001 |
|  | VCT | 2903 (38.2) | 1868 (35.5) | 1035 (44.4) |  |
|  | Inpatient | 98 (1.3) | 53 (1.0) | 45 (1.9) |  |
|  | Other | 716 (9.4) | 502 (9.5) | 214 (9.2) |  |
|  | Unknown | 3044 (40.0) | 2108 (40.0) | 936 (40.1) |  |
| Marital status | Single | 1174 (15.4) | 836 (15.9) | 338 (14.5) | <0.001 |
|  | Married/in union | 1588 (20.9) | 898 (17.0) | 690 (29.6) |  |
|  | Living with partner | 374 (4.9) | 271 (5.1) | 103 (4.4) |  |
|  | Divorced | 339 (4.5) | 283 (5.4) | 56 (2.4) |  |
|  | Widowed | 650 (8.6) | 580 (11.0) | 70 (3.0) |  |
|  | Unknow/Missing | 3476 (45.7) | 2400 (45.6) | 1076 (46.1) |  |
| Weight for adults, Kg | Median (IQR) | 59(52,67) | 58(51,66) | 60(54,67) | <0.001 |
|  | Missing n (%) | 1228(16.2) | 833(15.8) | 395(16.9) |  |
| Height for adults, CM | Median (IQR) | 162(157,169) | 160(155,165) | 170(165,175) | <0.001 |
|  | Missing n (%) | 1311(17.2) | 867(16.5) | 444(19.0) |  |
| BMI | Median (IQR) | 22.22(19.92,24.97) | 22.83(20.31,25.71) | 21.11(19.27,23.37) | <0.001 |
|  | Missing n (%) | 2379(31.3) | 1603(30.4) | 776(33.3) |  |
| CD4 count (cells/uL) | Median (IQR) | 361(205,543) | 392(232,584) | 293(160,454) | <0.001 |
|  | <100 | 549 (7.2) | 323 (6.1) | 226 (9.7) | <0.001 |
|  | 100-199 | 684 (9.0) | 414 (7.9) | 270 (11.6) |  |
|  | 200-349 | 1241 (16.3) | 827 (15.7) | 414 (17.7) |  |
|  | >350 | 2653 (34.9) | 2030 (38.5) | 623 (26.7) |  |
|  | Missing | 2474 ( 32.5) | 1674 (31.8) | 800 ( 34.3) |  |
| WHO Stage at enrollment | Stage I | 2064 (27.2) | 1508 (28.6) | 556 (23.8) | <0.001 |
|  | Stage II | 1238 (16.3) | 867 (16.5) | 371 (15.9) |  |
|  | Stage III | 1650 (21.7) | 1104 (21.0) | 546 (23.4) |  |
|  | Stage IV | 465 (6.1) | 278 (5.3) | 187 (8.0) |  |
|  | Missing | 2184 (28.7) | 1511 (28.7) | 673 (28.8) |  |
| Country | Rwanda | 5159 ( 67.9) | 3492 (66.3) | 1667 (71.5) | <0.001 |
|  | Burundi | 290 (3.8) | 238 (4.5) | 52 (2.2) |  |
|  | Cameroon | 1849 (24.3) | 1317 (25.0) | 532 (22.8) |  |
|  | DRC | 165 (2.2) | 121 (2.3) | 44 (1.9) |  |
|  | ROC | 138 (1.8) | 100 (1.9) | 38 (1.6) |  |
| Time Period | Year <= 2006 | 484 (6.4) | 326 (6.2) | 158 (6.8) | <0.001 |
|  | Year 2007 - 2010 | 1603 (21.1) | 1036 (19.7) | 567 (24.3) |  |
|  | Year 2011 - 2014 | 2566 (33.8) | 1854 (35.2) | 712 (30.5) |  |
|  | Year >= 2015 | 2948 (38.8) | 2052 (39.0) | 896 (38.4) |  |

\* more than 30 days prior to enrollment<sup>1</sup> By Chi-Square test

### Trends in measures of CD4 cell count and ART use prior to and after enrollment into care in CA-IeDEA

Before the WHO’s 2015 Treat All recommendations, CD4 cell count was an important criterion for determining eligibility for ART initiation. While CD4 testing is no longer required for determining eligibility for ART, pre-treatment CD4 testing is still recommended in order to identify patients with advanced disease who are at elevated risk for mortality. We examined trends in median CD4 cell count among all patients enrolling in HIV care at the participating clinics as well as ART use either prior to or after they enrolled in HIV care at the current site.

Among patients at participating CA-IeDEA sites, median CD4 cell count for all patients when they enrolled in care increased from 190 cells/uL in the period before 2005 to 334 cells/uL after 2017 (Figure 2). In 2005, the proportion of respondents who were already using ART before they enrolled in HIV care at the current CA-IeDEA participating site was less than 2% increasing to 18% in 2017. The proportion of patients who initiated ART within 30 days of enrollment in HIV care at the current CA-IeDEA site grew from 16% in 2005 to 75% after 2017.

**Fig. 2:**
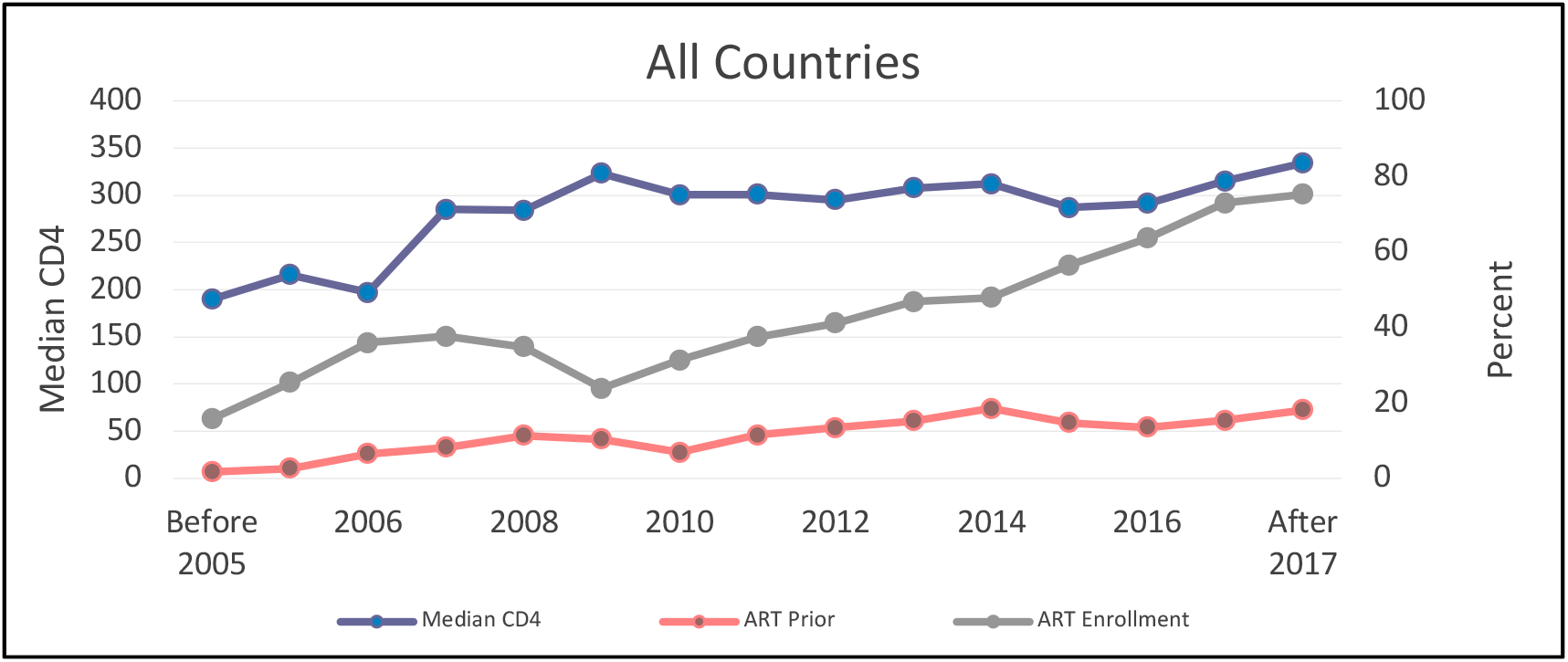
Trends in CD4 cell count and ART use before enrollment and after enrollment in HIV care among PLWH in all participating CA-IeDEA countries.

Disaggregated data by country is presented in Supplemental Figures S6-S10. In Burundi (Figure S6), recent median CD4 cell count at enrollment increased from 281 cells /uL in 2009 to 504 cells/uL in 2017 before a sharp decline to 372 cells/uL after 2017. About 5% of patients enrolling in HIV care at the participating CA-IeDEA site were already using ART before enrollment, but among those newly initiating ART, the proportion rose from about 10% in 2009 to 91% after 2017.

The trend in Cameroon (Figure S7) show median CD4 increase from 155 cells/uL in 2004 to 229 cells/uL in 2017 and falling to 177 cells/uL after 2017. The proportion of patients using ART prior to enrollment into CA-IeDEA participating clinic increased from 8% in 2004 to 22% in the period after 2017 while the proportion of patients initiating ART the first time when they enrolled in the CA-IeDEA participating site increased from 67% in 2004 to 74% after 2017.

In the DRC (Figure S8), median CD4 fluctuated from 327 cells/uL in 2005 to 295 cells/uL in 2006 and 375 cells/uL in 2007. Median CD4 cell count was highest in 2017 at 517 cells/uL. Similar fluctuations were observed with regard to ART use either prior to or after enrollment in HIV care at the participating clinic.

In the ROC (Figure S9), median CD 4 cell count among those enrolled in care fell from 221 cells/uL in the period prior to 2005 to as low as 142 cells/uL in 2005 before rising between 2006 and 2010 and then slightly from 2011 until 2015 and from 2016 onwards. Between 2005 and 2009, less than 4% of patients who enrolled in HIV at the current site were experienced ART users while the proportion of first-time users increased from 4% in 2005 to 51% in 2007 and again from 2010 onwards.

Trends in Rwanda were similar to those observed in the other countries (Figure S10). Recent median CD4 at enrollment increased in 2005 and then fell in 2006 before rising again and peaking at 457 cells/uL after 2017. The proportion of patients who were experienced ART users prior to enrollment was 2% in 2005 and 26% after 2017. The proportion of first time ART initiators increased 32% in 2005 to 67% after 2017.

## Discussion

In this study describing trends in demographic, clinical and immunological characteristics of the largest cohort of adult PLWH entering care and initiating ART in Central Africa from 2004-2018, we found increases in baseline CD4 cell count among patients enrolling in HIV care in the participating CA-IeDEA sites and in the proportion of first time ART users within 30 days of entry into care at participating sites in Burundi, Cameroon, DRC, ROC and Rwanda.

These findings shed light on important demographic, clinical and immunological characteristics of patients entering HIV care and those initiating ART, and also highlight the progress and challenges in early HIV diagnosis, and ART initiation that are critical to the universal goal to eliminate the virus by 2030. More importantly, our findings can inform effective programs and/or policies to improve long term outcomes and quality of life and substantially reduce mortality among PLWH living in Central Africa, particularly in the era of universal ART use. ^**11**^

Women comprised most of patients who enrolled in care and initiated ART within 30 days of enrollment in our CA-IeDEA cohort. While other studies^**12, 13**^ reported that HIV prevalence in SSA is higher among women than men, this discrepancy may reflect the fact that women were more likely to be tested for HIV earlier than men because of contacts with antenatal care during pregnancy.^**13**^ The higher proportion of women represented in the CA-IeDEA cohort may partly reflect disparities in testing, but may also reflect gender disparities and higher risk of transmission in women and perhaps, higher mortality rates in men.^**14**^

Before the WHO’s 2015 recommendation of universal HIV treatment for all PLWH, also known as “Treat All”, CD4 cell count, and WHO staging were the most commonly used criteria for determining treatment eligibility.^**15**^ Our findings suggest that at that time information on CD4 cell count and WHO staging was missing for a substantial proportion of patients, with likely implications for delayed timing of ART initiation. While there were differences between countries and between women and men, the low median CD4 cell counts at entry into care in the participating CA-IeDEA site suggests that PLWH were being diagnosed late and that they were likely entering care with advanced disease, particularly in the pre-Treat All era. In the post Treat All era, rising median CD4 cell counts suggests that patients were entering HIV care earlier, although it is worth highlighting that even after CD4 eligibility thresholds were eliminated, median CD4 cell count at entry into care remained less than 500 cells/uL. We also observed sex differences across countries with women having higher CD4 cell count at enrollment into HIV care at the participating CA-IeDEA site and ART initiation than men, perhaps reflecting women’s earlier access to HIV testing and diagnosis through antenatal care in PMTCT clinics.

Additionally, the fact that median CD4 count was quite low in the pre-treat all era suggests that patients may not have been accessing treatment early enough and may have been severely immune-compromised by the time treatment is initiated. Analysis of median CD4 count by country shows that patients in Rwanda had the highest median CD4 cell count when they enrolled in care in our participating clinics, which is higher than patients in Burundi, Cameroon, DR Congo and ROC.

Over the time period focused on in this study, there were changes in recommended criteria for ART initiation among PLWH, especially more recently, the Treat All guidelines that recommended immediate ART initiation upon HIV diagnosis regardless of clinical and immunological parameters.^**16**^ Arguably, changes in treatment guidelines may have contributed to the increased proportion of those initiating immediate ART use after entry into care at the participating clinics. Our finding show that ART initiation increased dramatically for PLWH enrolled from 2015 onwards, coinciding with the existence/implementation of the Treat All guidelines.

HIV Voluntary counseling and testing (HIV-VCT) is an important gateway to knowing one’s HIV status. VCT is beneficial for the individual by ensuring they know their HIV status and are able to take preventive actions against HIV acquisition or transmission to others. Additionally, knowledge of HIV seropositive status increases the likelihood of early engagement in care, facilitates timely ART initiation and confer protective benefits on the entire population.^**17**^ Consistent with evidence from other studies^**18**^, we found VCT clinics as an important route of entry into HIV care for a large proportion of patients in our cohort and this may have contributed to the rapid initiation of ART within 30 days of enrollment.

This descriptive study utilized data from 5 countries to describe the demographic, clinical and immunological characteristics of PLWH in Burundi, Cameroon, Democratic Republic of Congo, Republic of Congo and Rwanda. This combined data set that examined trends in demographic, clinical and immunological characteristics over a 14-year period in the largest cohort of adult PLWH entering care and initiating ART in Central Africa is a strength of the study. Nonetheless, there are limitations, which warrant caution in interpretation of the findings reported here. First, we highlight the different sample sizes of the cohort in each participating country. That nearly 4 in 10 CA-IeDEA patients were from Rwanda may have skewed the results described here. Second, missing data on weight, height, CD4 cell count and WHO staging in many clinics impacted the findings reported in this study. Additionally, the potential non-representativeness of the selected clinics and their concentration in largely urban and peri-urban settings is another limitation as clinics in urban and semi-urban settings do not necessarily represent all health facilities, especially those in rural settings providing HIV care in all countries. In addition, the participating clinics may be better resourced to scale up patient enrollment in care and treatment initiation, which may not reflect the disparity between urban/peri-urban and rural health facilities.

Despite these limitations, the CA-IeDEA as a cohort study offers an opportunity for further longitudinal analysis of the socio-contextual and health system factors that facilitate or hinder timely HIV diagnosis, enrollment in care, ART initiation among PLWH in the region. This is critical because Central Africa continues to lag behind other regions in curtailing the spread of HIV. For example, in-depth analyses are needed to examine trends in long-term outcomes among PLWH in this region so as to generate evidence to inform programs and policies that addresses the circumstances that are unique to each country and the region.

## Conclusion

Trends in demographic, clinical and immunological characteristics of patients entering into HIV care and subsequently initiating ART in the participating sites in Burundi, Cameroon, DRC, ROC and Rwanda highlight improvements over time among those accessing care in this cohort. Although progress is being made especially with improvements in CD4 cell count and the number of patients initiating ART soon after enrolling in care, gaps still remain with scaling up access to care for PLWH who may not be accessing care. Thus, implementing contextually relevant program and policy strategies to address gaps that remain is critical.

## Data Availability

Data for the Central Africa International Epidemiology to Evaluate AIDS (CA-IeDEA) is available upon request.

## Funding acknowledgement

Research reported in this publication was supported by the National Institutes of Health’s National Institute of Allergy and Infectious Diseases (NIAID), the *Eunice Kennedy Shriver* National Institute of Child Health & Human Development (NICHD), the National Cancer Institute (NCI), the National Institute on Drug Abuse (NIDA), the National Heart, Lung, and Blood Institute (NHLBI), the National Institute on Alcohol Abuse and Alcoholism (NIAAA), the National Institute of Diabetes and Digestive and Kidney Diseases (NIDDK), the Fogarty International Center (FIC), the National Library of Medicine (NLM), and the Office of the Director (OD) under Award Number U01AI096299 (Central Africa-IeDEA). The content is solely the responsibility of the authors and does not necessarily represent the official views of the National Institutes of Health.

## Site investigators and cohorts

Nimbona Pélagie, Association Nationale de Soutien aux Séropositifs et Malade du Sida (ANSS), Burundi; Patrick Gateretse, Jeanine Munezero, Valentin Nitereka, Théodore Niyongabo, Christelle Twizere, Centre National de Reference en Matière de VIH/SIDA, Burundi; Hélène Bukuru, Thierry Nahimana, Centre Hospitalo-Universitaire de Kamenge (CHUK), Burundi; Elysée Baransaka, Patrice Barasukana, Eugene Kabanda, Martin Manirakiza, François Ndikumwenayo, CHUK/Burundi National University, Burundi; Jérémie Biziragusenyuka, Ange Marie Michelline Munezero, Hospital Prince Régent Charles (HPRC), Burundi; Tabeyang Mbuh, Kinge Thompson Njie, Edmond Tchassem, Kien-Atsu Tsi, Bamenda Hospital, Cameroon; Rogers Ajeh, Mark Benwi, Marc Lionel Ngamani, Victorine Nkome, Falone Sandjong, Clinical Research Education and Consultancy (CRENC), Cameroon; Anastase Dzudie, CRENC and Douala General Hospital, Cameroon; Akindeh Mbuh, CRENC and University of Yaoundé, Cameroon; Djenabou Amadou, Amadou Dodo Balkissou, Eric Ngassam, Eric Walter Pefura Yone, Jamot Hospital, Cameroon; Alice Ndelle Ewanoge, Norbert Fuhngwa, Ernestine Kendowo, Chris Moki, Denis Nsame Nforniwe, Limbe Regional Hospital, Cameroon; Catherine Akele, Faustin Kitetele, Patricia Lelo, Martine Tabala, Kalembelembe Pediatric Hospital, Democratic Republic of Congo; Emile Wemakoy Okitolonda, Cherubin Ekembe, Kinshasa School of Public Health, Democratic Republic of Congo; Merlin Diafouka, Martin Herbas Ekat, Dominique Mahambou Nsonde, CTA Brazzaville, Republic of Congo; Adolphe Mafou, CTA Pointe-Noire, Republic of Congo; Nicole Ayinkamiye, Jules Igirimbabazi, Bethsaida Health Center, Rwanda; Emmanuel Ndamijimana, Providance Uwineza, Busanza Health Center, Rwanda; Emmanuel Habarurema, Marie Luise Nyiraneza, Gahanga Health Center, Rwanda; Dorothee Mukamusana, Liliane Tuyisenge, Gikondo Health Center, Rwanda; Catherine Kankindi, Christian Shyaka, Kabuga Health Center, Rwanda; Marie Grace Ingabire, Bonheur Uwakijijwe, Kicukiro Health Center, Rwanda; Jules Ndumuhire, Marie Goretti Nyirabahutu, Masaka Health Center, Rwanda; Yvette Ndoli, Oliver Uwamahoro, Nyarugunga Health Center, Rwanda; Ribakare Muhayimpundu, Sabin Nsanzimana, Eric Remera, Esperance Umumararungu, Rwanda Biomedical Center, Rwanda; Lydia Busingye, Alex M Butera, Josephine Gasana, Thierry Habiryayo, Charles Ingabire, Jules Kabahizi, Jean Chrysostome Kagimbana, Faustin Kanyabwisha, Gallican Kubwimana, Benjamin Muhoza, Athanase Munyaneza, Gad Murenzi, Francoise Musabyimana, Francine Mwiza, Gallican Nshogoza Rwibasira, Jean d’Amour Sinayobye, Patrick Tuyisenge, Rwanda Military Hospital, Rwanda; Chantal Benekigeri, Jacqueline Musaninyange, WE-ACTx Health Center, Rwanda.

## Coordinating and Data Centers

Adebola Adedimeji, Kathryn Anastos, Madeline Dilorenzo, Lynn Murchison, Jonathan Ross, Marcel Yotebieng, Albert Einstein College of Medicine, USA; Diane Addison, Ellen Brazier, Heidi Jones, Elizabeth Kelvin, Sarah Kulkarni, Denis Nash, Matthew Romo, Olga Tymejczyk, Institute for Implementation Science in Population Health, Graduate School of Public Health and Health Policy, City University of New York (CUNY), USA; Batya Elul, Columbia University, USA; Xiatao Cai, Allan Dong, Don Hoover, Hae-Young Kim, Chunshan Li, Qiuhu Shi, Data Solutions, USA; Robert Agler, Kathryn Lancaster, The Ohio State University, USA; Mark Kuniholm, University at Albany, State University of New York, USA; Andrew Edmonds, Angela Parcesepe, Jess Edwards, University of North Carolina at Chapel Hill, USA; Olivia Keiser, University of Geneva; Stephany Duda; Vanderbilt University School of Medicine, USA; April Kimmel, Virginia Commonwealth University School of Medicine, USA.

## Supplementary Materials

**Figure. S1:**
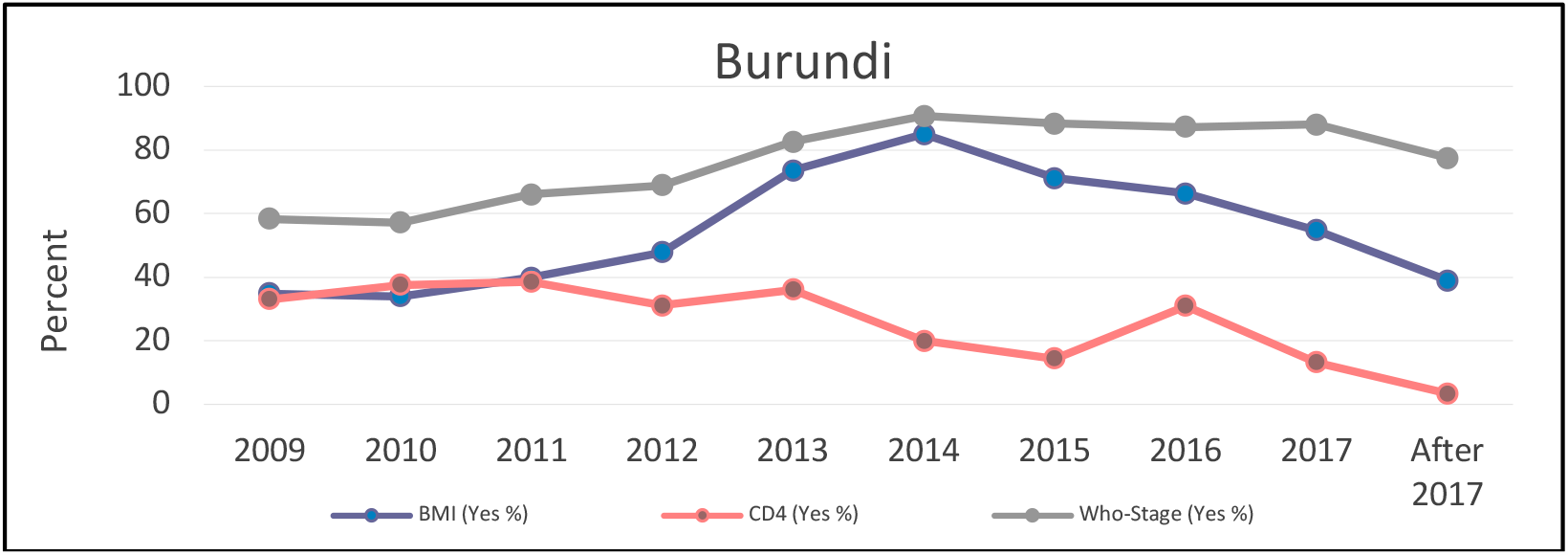
Trends in proportions having a recent measure for BMI, CD4 cell count and WHO stage in the Burundi CA-IeDEA cohort.

**Figure. S2:**
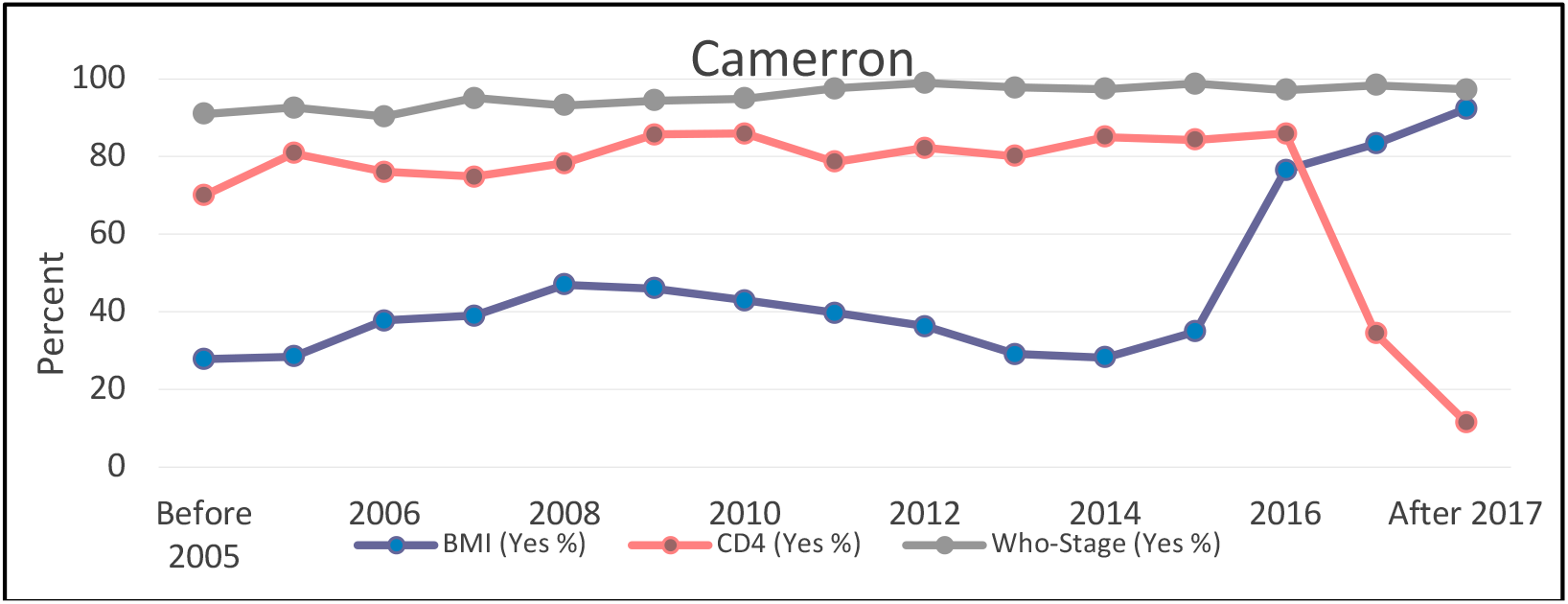
Trends in proportions having a recent measure for BMI, CD4 cell count and WHO stage in the Cameroon CA-IeDEA cohort.

**Figure S3:**
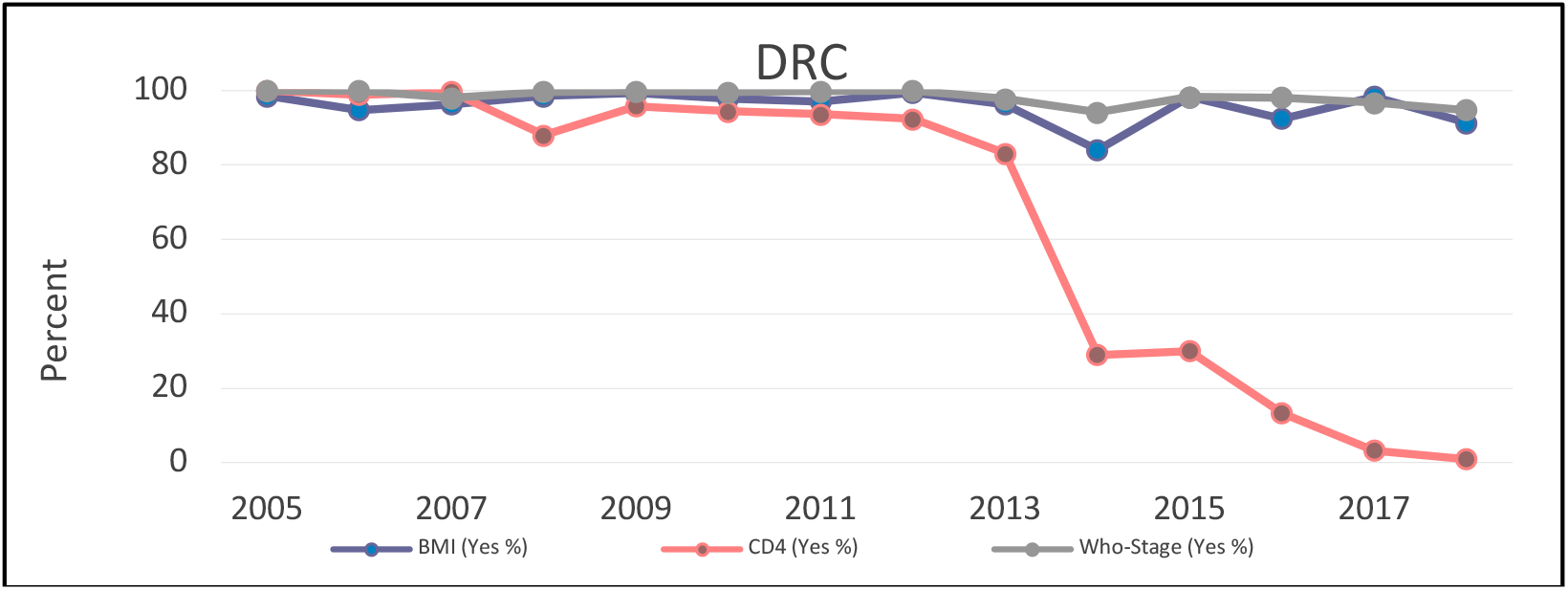
Trends in proportions having a recent measure for BMI, CD4 cell count and WHO stage in the DRC CA-IeDEA cohort.

**Fig. S4:**
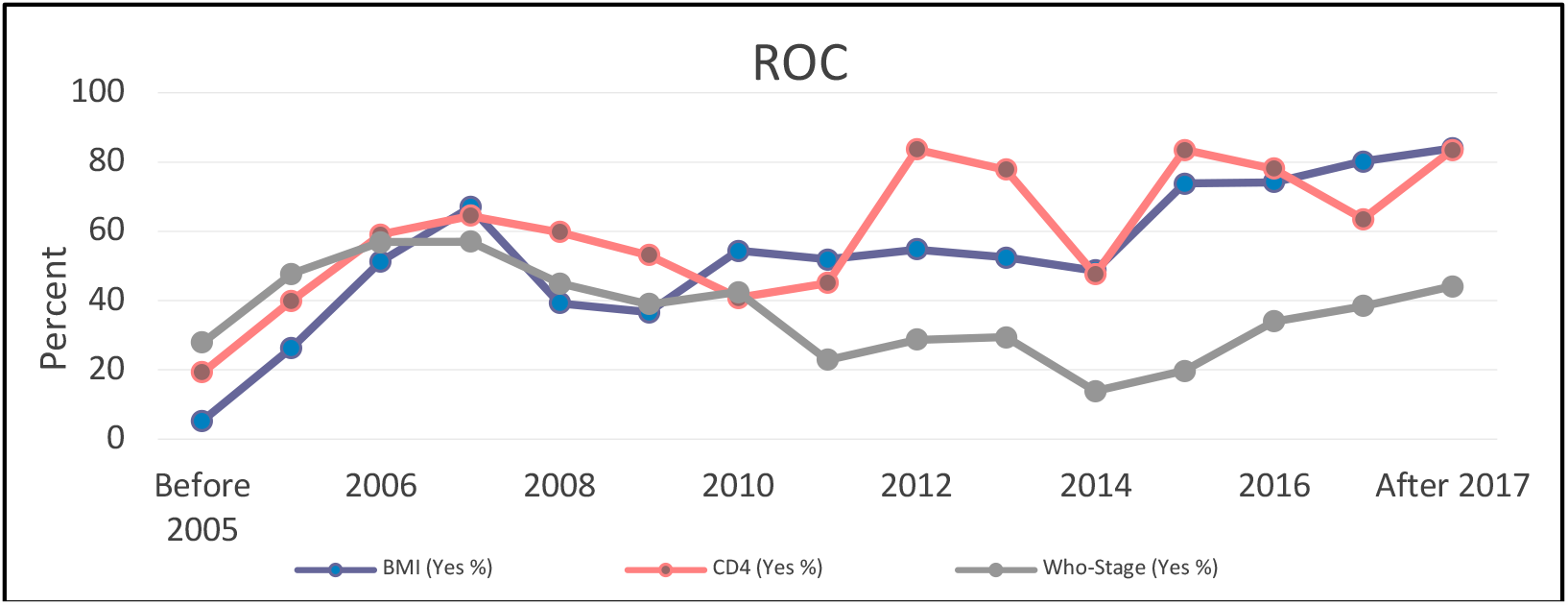
Trends in proportions having a recent measure for BMI, CD4 cell count and WHO stage in the ROC CA-IeDEA cohort.

**Fig. S5:**
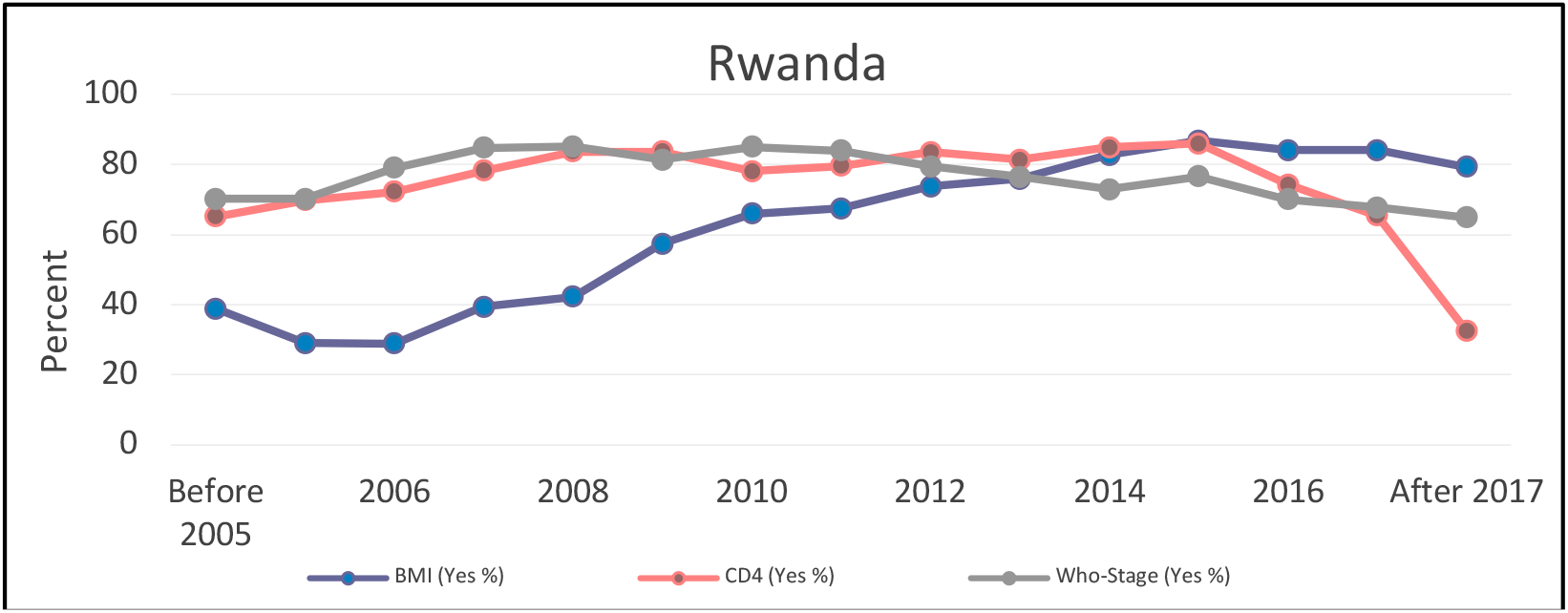
Trends in having a recent measure for BMI, CD4 cell count and WHO stage at enrollment in HIV care in the Rwanda CA-IeDEA cohort.

**Fig. S6:**
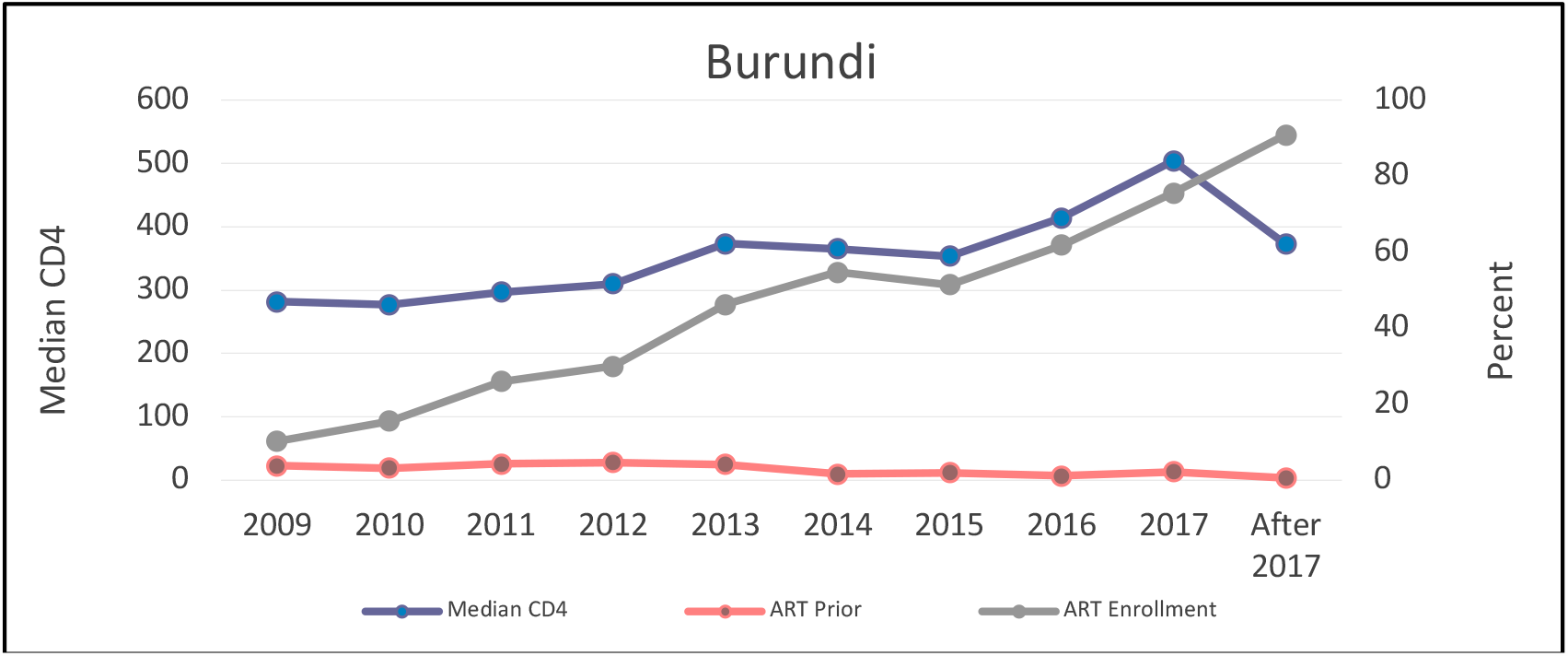
Trends in CD4 cell count and ART use before enrollment and after enrollment in HIV care among PLWH in Burundi CA-IeDEA cohort.

**Fig. S7:**
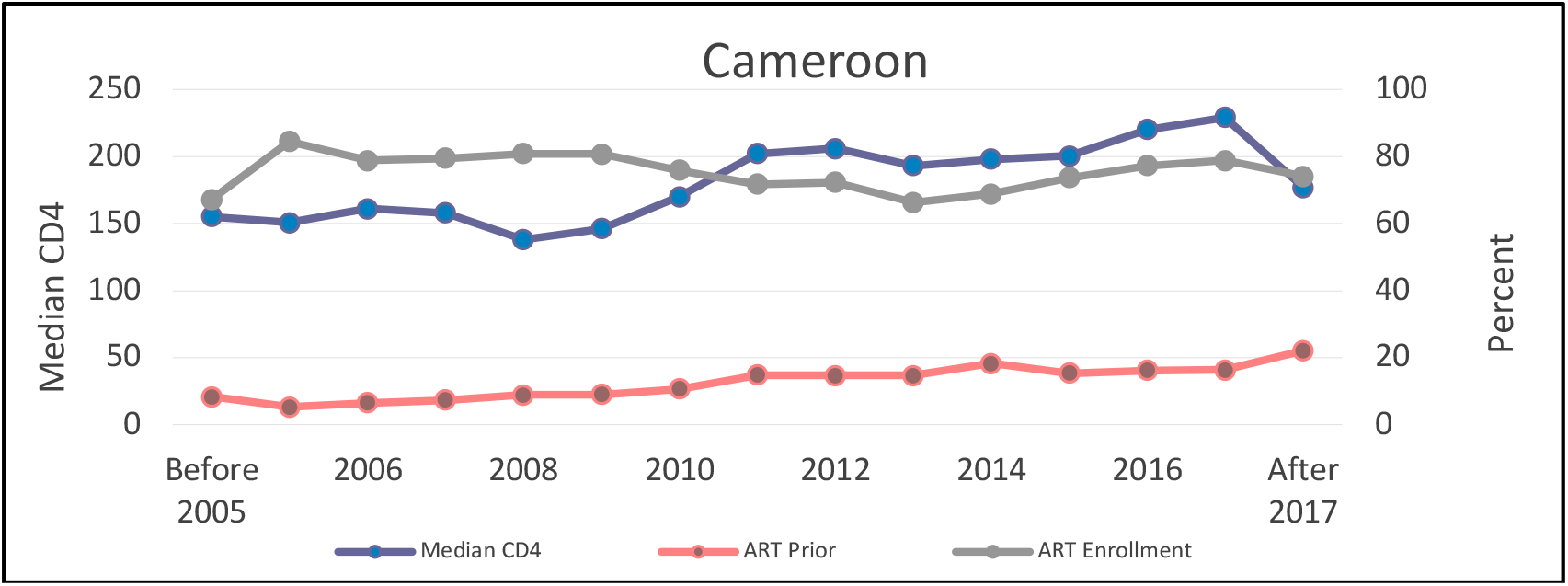
Trends in CD4 cell count and ART use before enrollment and after enrollment in HIV care among PLWH in Cameroon CA-IeDEA cohort.

**Fig. S8:**
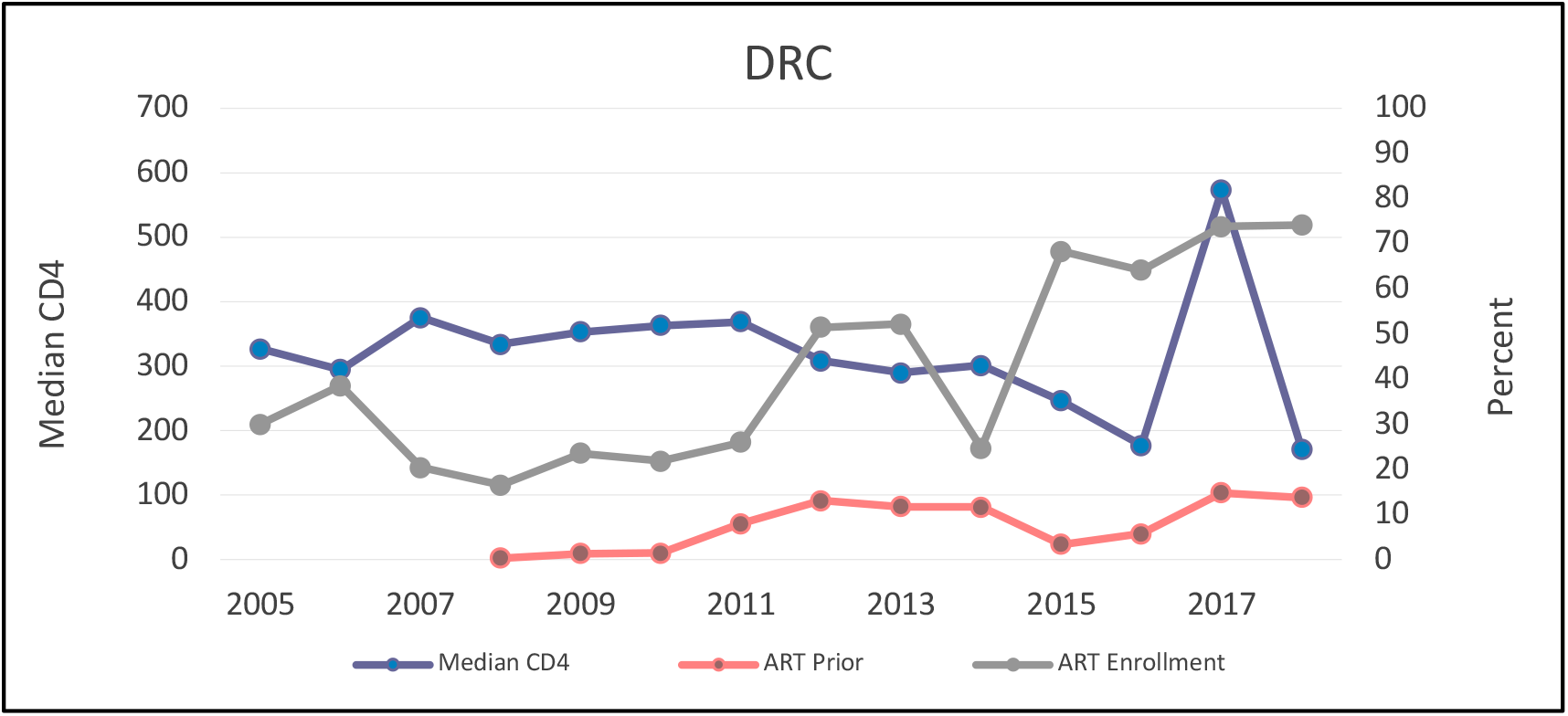
Trends in CD4 cell count and ART use before enrollment and after enrollment in HIV care among PLWH in DRC CA-IeDEA cohort.

**Fig. S9:**
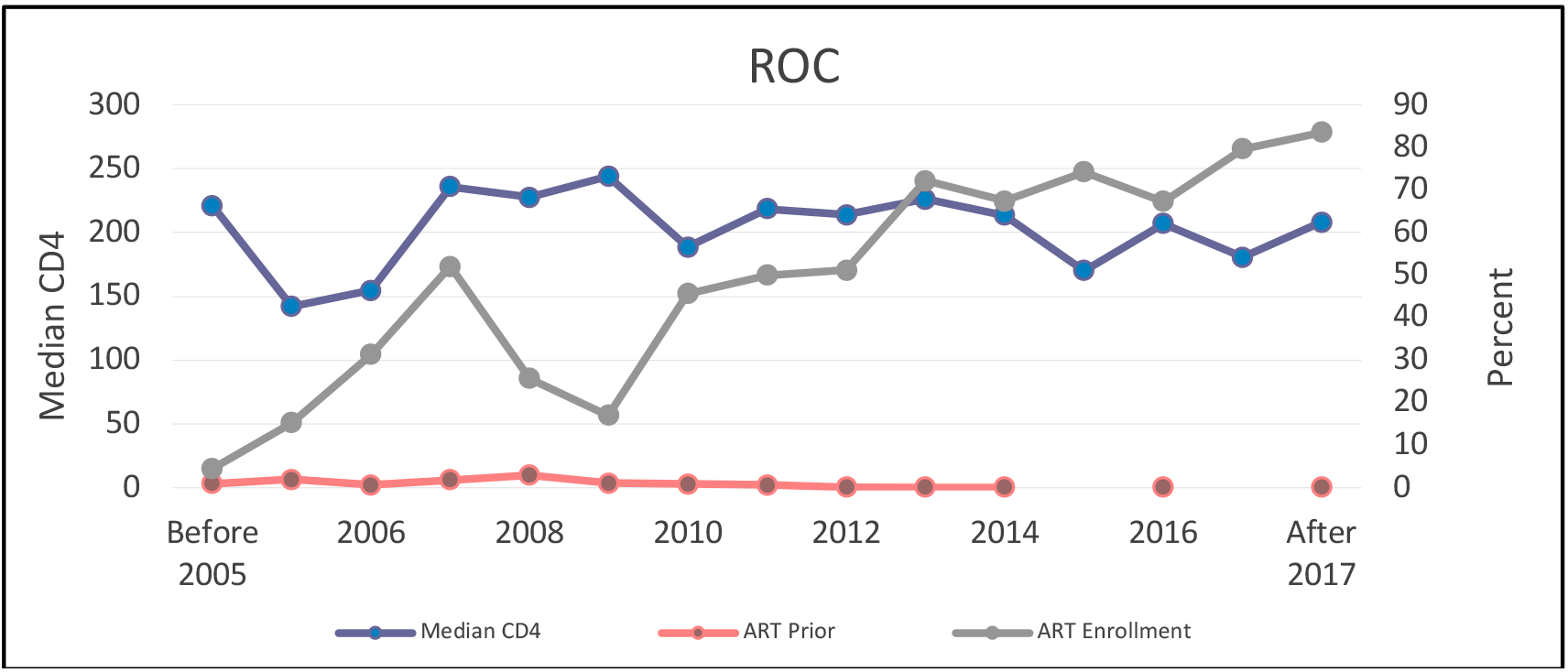
Trends in CD4 cell count and ART use before enrollment and after enrollment in HIV care among PLWH in ROC CA-IeDEA cohort.

**Fig. S10:**
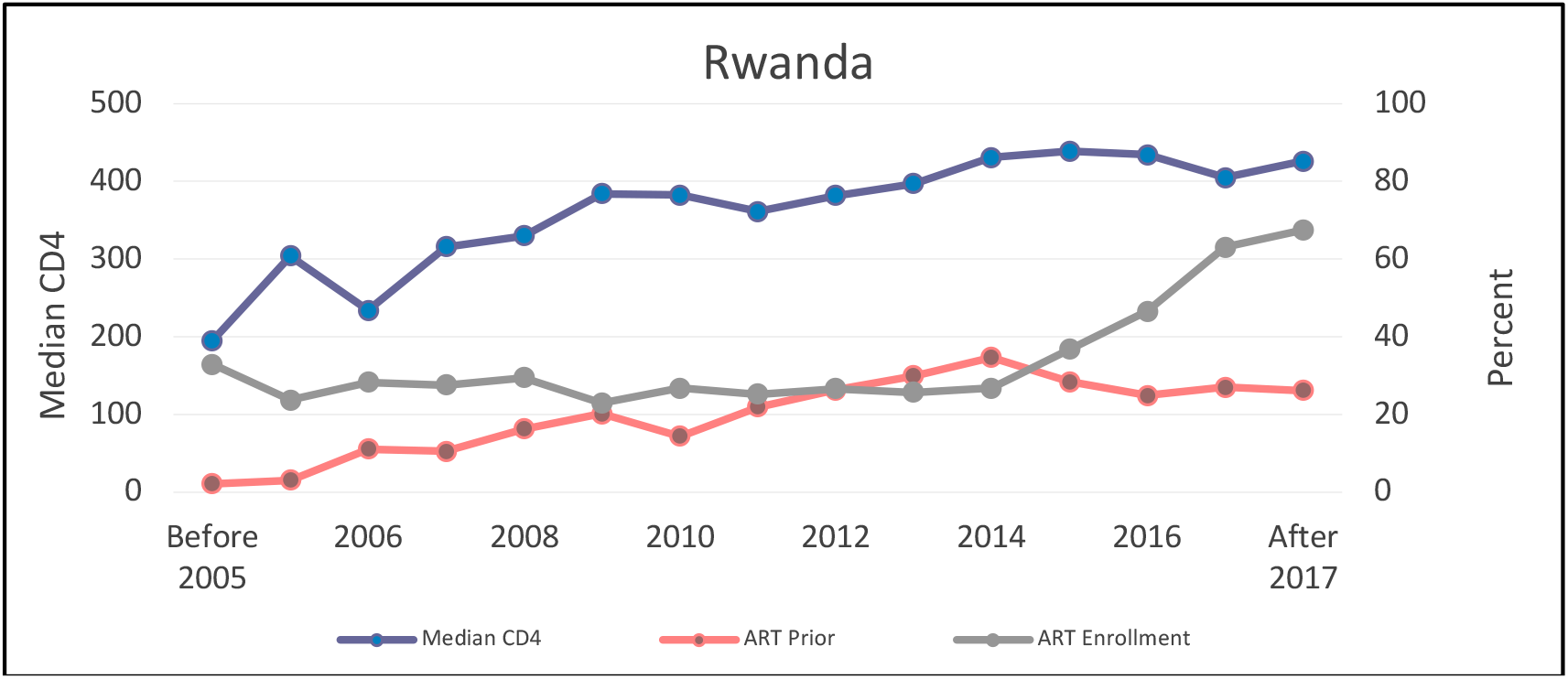
Trends in CD4 cell count and ART use before enrollment and after enrollment in HIV care among PLWH in Rwanda CA-IeDEA cohort.

